# Attention-based whole-slide image compression achieves pathologist-level pre-screening of multi-organ routine histopathology biopsies

**DOI:** 10.1101/2024.12.17.24319180

**Authors:** Witali Aswolinskiy, Rachel S van der Post, Michiel Simons, Enrico Munari, Michela Campora, Carla Baronchelli, Laura Ardighieri, Simona Vatrano, Iris Nagtegaal, Jeroen van der Laak, Francesco Ciompi

## Abstract

Screening programs for early detection of cancer such as colorectal and cervical cancer have led to an increased demand for histopathological analysis of biopsies. Advanced image analysis with Deep Learning has shown the potential to automate cancer detection in digital pathology whole-slide images. Particularly, techniques of weakly supervised learning can achieve whole-slide image classification without the need for tedious, manual annotations, using only slide-level labels. Here, we used data from n=12,580 whole-slide images from n=9,141 tissue blocks to train and validate a deep learning approach based on Neural Image Compression with Attention (NIC-A) and show how it can be leveraged to pre-screen (pre)malignant lesions in colorectal and cervical biopsies and to analyze duodenal biopsies for celiac disease. Our NIC-A classifies normal tissue, low-grade dysplasia, high-grade dysplasia and cancer in colon and uterine cervix, and identifies celiac disease in duodenal biopsies. We validated NIC-A for colon and cervix against a panel of four and three pathologists, respectively, on cohorts from two European centers. We show that the proposed approach reaches pathologist-level performance at detecting and classifying abnormalities, suggesting its potential to assist pathologists in pre-screening workflows by reducing workload in digital pathology routine diagnostics.

## Introduction

Population screening programs are producing an increasing number of histopathology tissue samples. Visual inspection, diagnosis and reporting of digital pathology whole slide images (WSI) of these large numbers of biopsies is a time-consuming process, leading to increased workloads and pressure on pathologists [1, 2], possibly prone to intra- and inter-observer variability. Furthermore, the screening-derived nature of these data results in the vast majority of tissue samples being clinically non informative, suggesting the need for methods to optimize diagnostic procedures.

In recent years, artificial intelligence (AI) algorithms based on deep learning (DL) have been proposed to address a variety of digital histopathology tasks, including detection of breast cancer metastases in lymph nodes, prostate and breast cancer grading [3].

In most cases, development of DL models is based on so-called supervised learning, trained using patch-(i.e., small portions of digital pathology images) or pixel-level manual annotations of different tissue types. While this type of approach can achieve high performance, it is grounded on a very tedious and time-consuming process [4]. As an alternative, weakly-supervised learning has been proposed [5] as a learning paradigm to train DL algorithms by solely using a slide- or case-level label, which can be extracted from readily available pathology reports, without the need for manual annotations. In recent years, researchers have equipped these methods with a so-called attention mechanism [6, 7], which allows DL algorithms to learn visual cues focusing on regions of the WSI relevant for the task at hand, such as tumor detection, improving both model’s predictive performance and interpretability. Trained with enough data samples, these approaches have shown high performance on numerous tasks [8, 9]. While different variations of this approach have been already applied to colorectal [10], cervical [11], and duodenal biopsies [12], a single framework capable of tackling multiple applications has not been studied so far. Furthermore, to the best of our knowledge, the inter-observer variability of pathologists on these diagnostic tasks has not been studied so far, limiting insights in the potential value of these algorithms if deployed in clinical practice.

In this study, we present a unified, weakly supervised framework to develop DL models for detection and classification of abnormalities in colorectal and cervical biopsies, targeting clinically relevant classes such as low-grade and high-grade dysplasia, and invasive carcinoma, as well as celiac disease (CD) in duodenal biopsies. We use the previously developed Neural Image Compression framework [13], equipped with an Attention mechanism [6, 7], which we refer to as NIC-A. As backbone for NIC-A, we used “foundational models”, artificial neural networks pre-trained to extract (i.e., encode) meaningful features from a large set of digital pathology images. We also make NIC-A compatible with the diagnostic setting when multiple slides per block are available by proposing a novel algorithm to “pack” multiple WSI into a “macro WSI”, facilitating both the training procedure when using block level labels extracted from pathology reports and simplifying viewing of the multiple tissue sections. Finally, we evaluate our approach for colon and cervix against a panel of experienced pathologists showing its potential to facilitate the diagnostic workflows.

## Material and Methods

### Study setup

We investigate the performance of NIC-A for sub-typing colorectal, cervical and duodenal biopsies into clinically relevant categories. To evaluate our models, we performed reader studies with a panel of four pathologists for colon and three pathologists for cervix biopsy diagnosis. We report the inter-observer agreement on these two use cases, and compare the best DL models with pathologists for sub-typing colon and cervix biopsies.

### Data

In this section, we describe the data used for training and validation of the proposed NIC-A approach for the classification of colon, cervix and duodenal biopsies. Additional data was used for evaluation of the performance of NIC-A in comparison with a panel of pathologists in reader studies. Table 1 provides an overview of the used data. Data was collected from the pathology archive of the Radboud University Medical Center (Radboudumc, Nijmegen, Netherlands) and from the Azienda Ospedaliera Cannizaro and Gravina Hospital Caltagirone (ASP, Catania, Italy). All slides from Radboudumc were scanned with a Pannoramic 1000 DX (3DHistech) at a spatial resolution of 0.24 μm/px; all slides from ASP were scanned with a Aperio AT2 (Leica Biosystems) at a spatial resolution of 0.24 μm/px.

**Table 1:** Data overview for the colon, cervix and celiac cohorts: Numbers of blocks (packed slides) from Radboudumc and individual slides from ASP per subset. Radboudumc: Radboud University Medical Center; ASP: Azienda Ospedaliera Cannizzaro and Gravina Hospital Caltagirone.

| Tissue type | Source | Subset | Label |  |  |  |  | Total |
| --- | --- | --- | --- | --- | --- | --- | --- | --- |
|  |  |  | Normal | LGD | HGD | Cancer | Celiac |  |
| Colon | Radboud umc | all | 4945 | 1501 | 201 | 296 | - | 6943 |
|  |  | training | 3911 | 1189 | 146 | 222 | - | 5468 |
|  |  | validation | 1008 | 292 | 43 | 60 | - | 1403 |
|  |  | evaluation | 26 | 20 | 12 | 14 | - | 72 |
|  | ASP | evaluation | 14 | 35 | 16 | 11 | - | 76 |
| Cervix | Radboud umc | all | 424 | 179 | 493 | 106 | - | 1202 |
|  |  | training | 319 | 142 | 379 | 61 | - | 901 |
|  |  | validation | 79 | 31 | 104 | 16 | - | 230 |
|  |  | evaluation | 26 | 6 | 10 | 29 | - | 71 |
|  | ASP | evaluation | 17 | 13 | 14 | 30 | - | 74 |
| Duodenum | Radboud umc | all | 571 | - | - | - | 425 | 996 |
|  |  | training | 414 | - | - | - | 298 | 712 |
|  |  | validation | 107 | - | - | - | 77 | 184 |
|  |  | evaluation | 50 | - | - | - | 50 | 100 |

#### Colon polyps and biopsies data

For the colon cohort, we retrospectively collected clinical cases from Radboudumc diagnosed between 2000 and 2009. Slide selection was based on an automated analysis of the diagnostic reports, excluding slides containing ileum, duodenum, stomach or small intestine tissue. The final collection consisted of 9,237 colorectal slides cut from 6,943 paraffin blocks of 3670 cases. Since the pathology reports referenced the paraffin blocks rather than individual slides, we combined WSI originating from the same block (see section ‘Slide packing’ for details). From this set, we used 6,871 slides to develop our models, and 72 slides from patients not included in the development set for evaluation. Furthermore, we included 76 slides from ASP with associated reports per slide for evaluation. Clinical conclusions in pathology reports were labeled by native-speakers (i.e., Italian for ASP, Dutch for Radboudumc reports) research assistants into the classes i) normal, ii) hyperplastic polyps, iii) low-grade dysplasia (LGD), iv) high-grade dysplasia (HGD) and v) cancer.

#### Uterine cervix cancer data

For the cervix cohort, we retrospectively collected cases diagnosed at Radboudumc between 2009 and 2020. Slide selection was based on an automated analysis of the diagnostic reports, excluding slides containing tissue from the endometrium and vagina. The resulting collection contained 2,312 slides cut from 1,272 paraffin blocks of 908 cases. Similar to the Radboudumc colon cohort, pathology reports referenced the paraffin blocks. From this set, we used 1,138 blocks for training and validation of NIC-A, and 71 blocks from different cases for evaluation. Additionally, we included 74 slides from ASP, with reports associated with the slides. Same as for colon, model training was done using only Radboudumc slides, while all slides from ASP were used for evaluation only. The reports were labeled into the classes: i) benign, ii) low-grade dysplasia (CIN1), iii) high-grade dysplasia (CIN2-3 + in-situ cancer) and iv) a combined class for invasive adenocarcinoma and squamous cell carcinoma (due to the small number of cancer cases).

#### Duodenal biopsies data

For the celiac disease (CD) cohort, we retrospectively collected 1,031 duodenal biopsy slides cut from 996 blocks of 961 Radboudumc patients (413 diagnosed with CD). 100 of these cases were used for internal evaluation. Slides from Radboudumc were scanned with a 3DHistech at 40x (0.25 microns pixel spacing) while slides from ASP were scanned with 3DHistech and Aperio scanners at 40x and 20x, respectively (0.25 and 0.5 microns per pixel spacing).

### Reader studies

To assess the inter-observer variability at the level of slide diagnosis, we conducted reader studies for the colorectal and cervical biopsy scoring via the web-based Grand Challenge platform [14].

The colon reader study consisted of 148 colorectal biopsies and polyps (72 from Radboudumc and 76 from ASP) independently scored by four pathologists (CvdP, EM, MC, CB). The pathologists could score cases as i) benign (normal or with hyperplastic or inflammatory polyps), with ii) LGD or iii) HGD in polyps and iv) adenocarcinoma. Furthermore, they could mark cases as ‘diagnosis not possible’ and provide a textual comment.

The cervix reader study was conducted in a similar fashion and contained 145 cervical biopsies (71 from Radboudumc and 74 from ASP), which were scored by three pathologists (MS, EM, LA). Comparable to colon, the choices were grouped in four categories: i) normal/benign (including reactive tissue), ii) LGD (including CIN1 and cylindrical atypia), iii) HGD (including CIN2, CIN3 and adenocarcinoma in-situ) and iv) cancer (including invasive squamous cell carcinoma and adenocarcinoma). The full list of choices in the reader studies is shown in the supplementary material.

The pathologists were from different institutes, and had different levels of expertise in diagnosis of colon and cervix biopsies. Each pathologist performed the study independently, blind to the results of the others. The reasons that were given for non-diagnosable slides included artifacts, poor quality, and necessity for immunohistochemistry or re-cut. In the set of results, we also considered the diagnosis from the original report as an additional reader.

At the end of each reader study, we derived a “reference standard” for each study from the collective knowledge of readers via majority voting, defined here as the same diagnosis achieved by three readers (including the original diagnosis from the report as one of the readers).

For cases where no reference standard could be obtained via majority voting (n=5 colon and n=32 cervix cases), two review meetings were organized. For colon, one pathologist from the reader study (CvdP) and one independent senior GI pathologist from Radboudumc (IN) reviewed the cases together. For cervix, all three participating pathologists met (virtually) and discussed the cases. For two colon and two cervix cases no consensus could be achieved. To evaluate the developed AI algorithms, we use the combination of the majority and review vote (referred here to as ‘Consensus’) as the reference standard (excluding the four non-diagnosable cases).

### Automated diagnosis via attention-based Neural Image Compression

We developed an approach to automate the diagnosis of biopsies using attention-based neural image compression. It consists of three steps (see Figure 1): ‘Packing’ tissue sections from several slides of a tissue block into a single slide, compressing (encoding) the slide using a pre-trained neural network and classifying the compressed slide. heatmap highlighting tissue patches relevant for the prediction result (relevance increases from blue to red).

**Figure 1:**
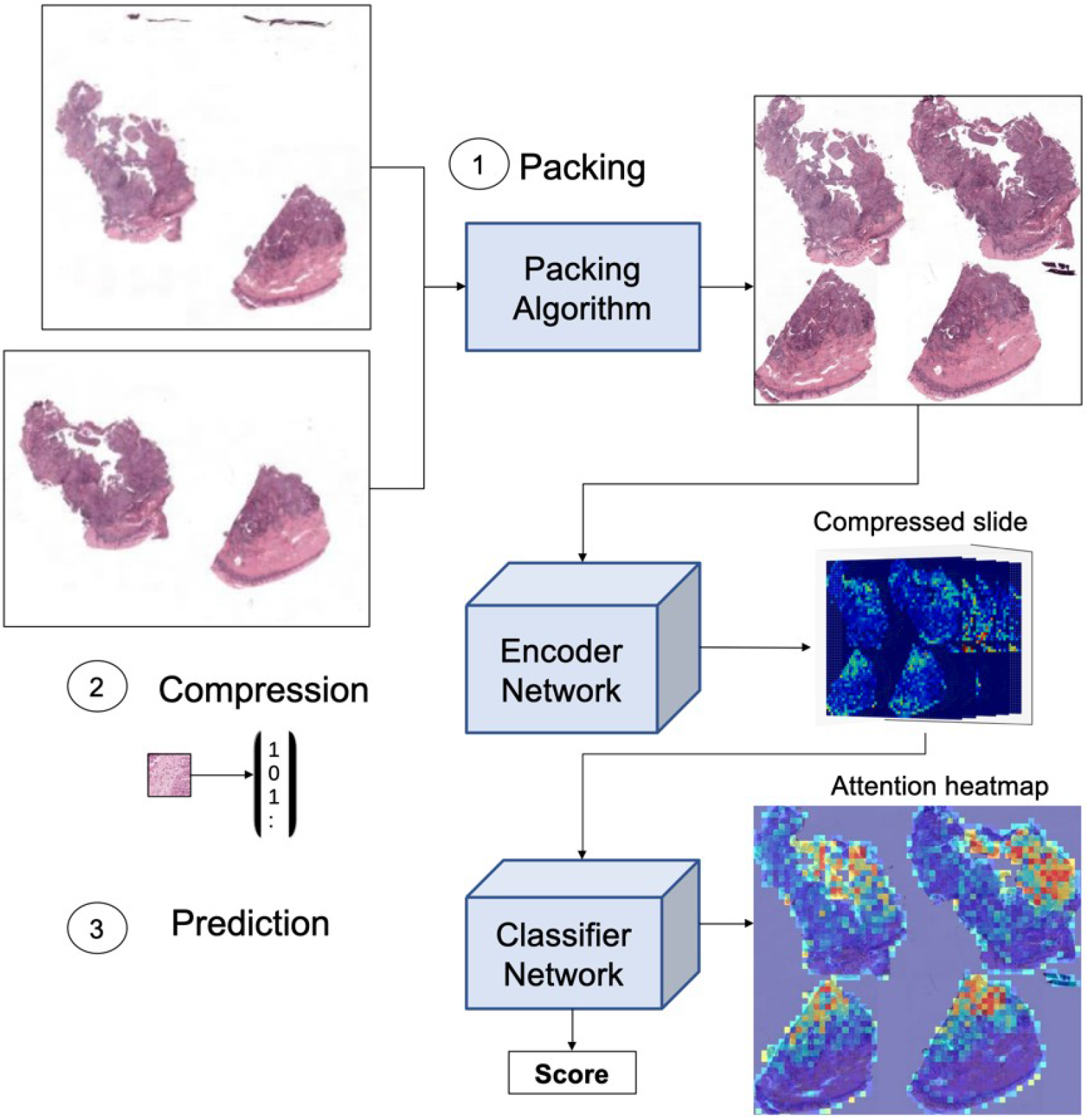
Method overview: First, the slides are packed, combining sections from slides of the same block. Then, the packed slides are compressed using pretrained foundational models (the shown neuron activations increase from blue to red). Finally, the encodings are classified with attention based neural image compression producing both a prediction score and an attention

#### Slide packing

For the Radboudumc cohorts, the diagnostic reports reference tissue blocks rather than individual slides. However, it may occur that only one fragment of a particular slide contains tumor cells while the others contain benign tissue irrelevant for the diagnosis, which poses a challenge for computer algorithms designed to learn from single WSIs. To address this, we developed a “WSI packing” procedure, which works in three steps. First, it collects all whole-slide images belonging to the same tissue block and identifies the tissue samples in those slides. Second, it separates the identified tissue (i.e., the foreground) from the rest of the slide (i.e., the background), and crops each foreground part using a bounding box that encapsulates the tissue. Third, it combines all tissue sections into a single new whole-slide image while minimizing the background area. To identify foreground regions, we apply a tissue segmentation model from previous work [15], and use “rectangle packing” [16, 17] to efficiently arrange these sections on the new image. This procedure not only addresses the challenge of the block-level labels but also reduces the file size of the resulting ‘packed’ images by removing the background area outside of the rectangles.

#### Encoders for Neural Image Compression

Following [6, 7, 13, 18], the packed slides are first processed using a pre-trained neural network, which acts as an encoder, receiving image tiles as input and producing for each a much smaller feature vector that encodes (i.e., describes) morphological features of the tissue as output. The quality of the compression and the amount of semantic and morphological information kept in the extracted features determines the performance on the downstream tasks. A common approach to design an encoder is to pre-train it on data different from the target application, often also belonging to a different data domain (e.g., natural images). Early studies showed that no single pre-training method consistently stands out among a set of pre-training techniques [8, 9], and that relevant pre-training approaches can be categorized in two groups: 1) supervised training using large datasets of natural images such as ImageNet [7] or histopathology data [19], and 2) unsupervised training on large unlabeled datasets of histopathology data [9].

In this work, we set out to identify the best foundational model to encode data for the applications at hand, evaluating and comparing a set of publicly available encoders among the state of the art at the time of designing this study, including early examples of histopathology foundation models.

We included three groups of publicly available models, built with various architectures and pretraining methods: 1) two convolutional encoders pretrained with supervision to solve an image classification task, one pretrained on ImageNet (ImageNet-Res50 [7]), producing feature vectors of size 1024, and one pretrained on 22 histopathology tasks (MTDP-Res50 [19]), producing feature vectors of size 2048; 2) two convolutional encoders pretrained on histopathology data via self-supervision: one pre-trained with the SimCLR approach [20] (HSSL-Res18 [21]), producing a feature vector of size 512, and one with the Barlow Twins approach [22] (Lunit-BT-Res50 [9]), producing a feature vector of size 2048; 3) three transformer-based encoders pretrained with self-supervision using the DINO approach [23] (HIPT256 [24] and Lunit-DINO16/8 [9]), producing a feature vector of size 384, where in HIPT we used the lowest-level transformer as encoder. For the Lunit transformer encoders we used patch size 224×2224 px and 256×256 px for the rest.

#### Attention-based classifier

After encoding, a classifier is trained to solve each sub-typing task, using as input the collection of the feature vectors organized spatially according to their position in the input slide, and the given diagnostic label as output target. Since we keep the spatial organization of the slide in the set of encoded features, following [13], we refer to it as the “compressed” whole-slide image. For the classifier network, a simple, attention-based architecture has been shown to yield state-of-the-art performance [6,7]. We use an equivalent architecture in our work, depicted in Suppl. Fig. 2, consisting of a convolution, followed by a multiplication with the attention values produced by the attention block and a final linear layer. The attention values from the attention block allow one to visualize where the network ‘looks at’ when classifying the slide. All convolutions have kernel sizes of 1, as increasing the kernel size did not improve performance in preliminary experiments.

**Figure 2:**
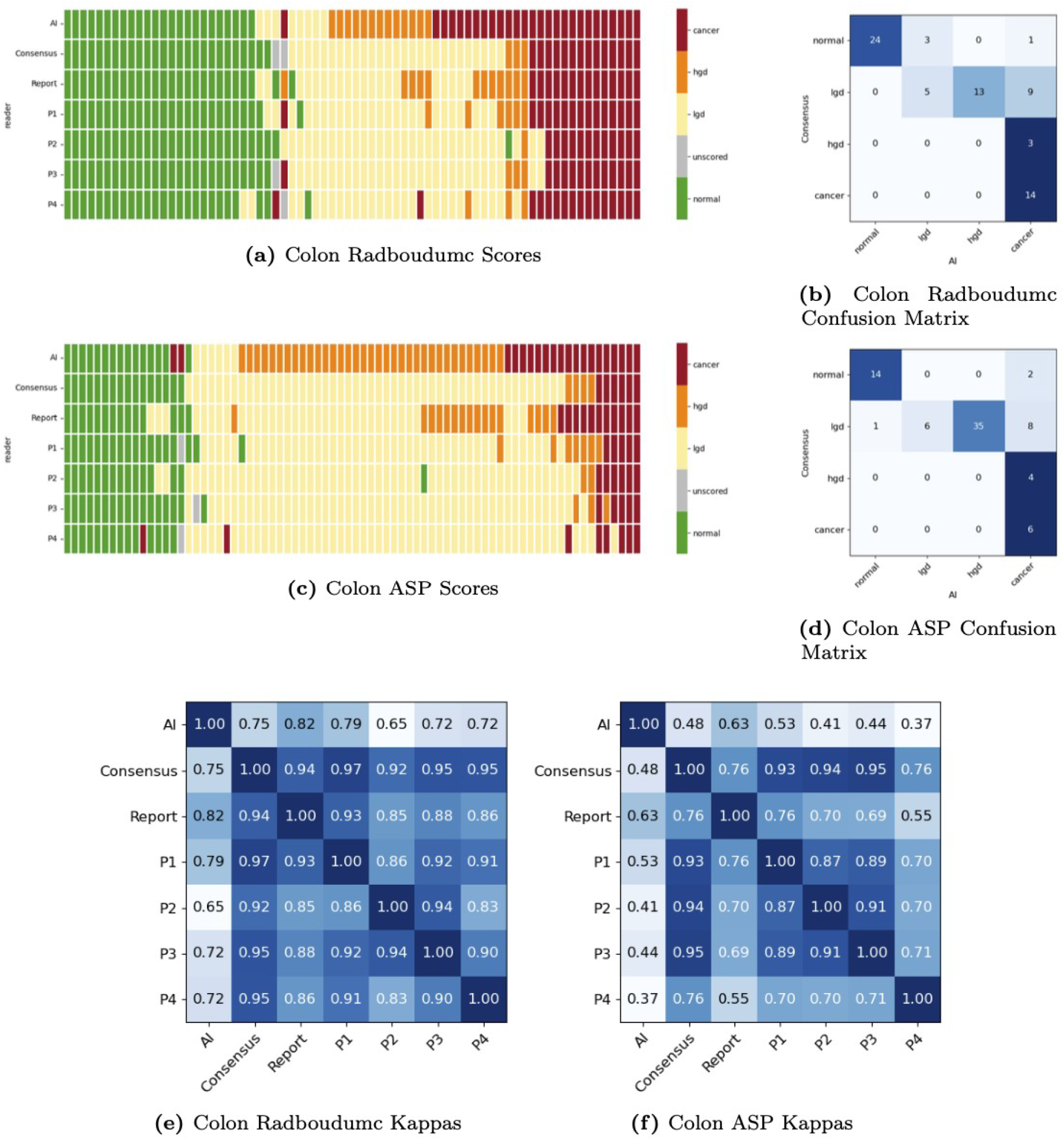
Colon reader study evaluation results. Slide scores (a,c), confusion matrices when compared to the consensus (b,d) and quadratic kappas for the inter-observer agreement (e,f) for the Radboudumc and ASP cohorts, respectively.

#### Diagnostic applications

Based on the same framework, we trained NIC-A models for the colon, cervix and celiac disease separately. In the rest of this paper, we refer to the NIC-A models for each application as NIC-A-colon, NIC-A-cervix and NIC-A-celiac.

In all cases, we first encoded each packed whole-slide image and then trained an attention-based (convolutional) neural network as a classifier for each of the diagnostic tasks. For celiac disease, we trained the classifier to predict celiac vs. non-celiac, using a binary cross entropy loss. For the diagnosis of colon and cervix biopsies, we trained the classifiers in a multi-class fashion with the classes “benign”, “LGD”, “HGD” and “cancer” using a cross entropy loss. Additionally, we added a binary cross-entropy loss for “benign” vs. “(pre)malignant” (grouping LGD, HGD and cancer together) to reflect the clinical importance of distinguishing benign from abnormal cases.

All networks were trained with class-balanced sampling for at most 150 epochs, stopping early after 25 epochs if there was no improvement in the average validation ROC-AUC (area under the receiver operating characteristic curve). We used the AdamW-optimizer with learning rate 1e-4 reduced by factor 10 if there was no improvement after 10 epochs. For each application, we trained three models with each encoder and selected the best one for the evaluation based on the internal validation ROC-AUC (see Table 1 for the data partition overview) .

## Results

We first report the results of the comparison of multiple compression encoders with performance metrics evaluated on the validation set. Based on these results, we selected the best performing method for each application, and compared them with the performance of a panel of pathologists on independent evaluation sets used in the reader studies.

### Selecting application-specific encoders

We compared a pool of pre-trained artificial neural network models including both convolutional neural networks and vision transformers for colon, cervix and celiac applications independently. Table 2 shows the per-application classification performance for each considered encoder (best models highlighted in bold). For colon, the best performance was achieved with a ResNet50 encoder pre-trained with self-supervision on histopathology data with Barlow Twins achieving an AUC=0.9693. For cervix, the best performance was achieved with a Vision Transformer pre-trained on histopathology data with DINO achieving AUC=0.8876. The self-supervised ResNet50 encoder was also in the top-3 with AUC=0.8761, performing only marginally worse than ViT. In general, we observed that most encoders achieved comparable AUC values for colon (AUC range 0.9542-0.9693) whereas for cervix the differences were more substantial (AUC range 0.7747-0.8876). For celiac disease, we observed that the best encoder was a ResNet50 pre-trained with full supervision using natural image data. However, for this application we observed that most models performed in a comparable way with most AUC ≥ 0.98, except for HIPT256 (AUC=0.9551). Based on these results, we selected the best encoders to be used in downstream analyses.

**Table 2:**
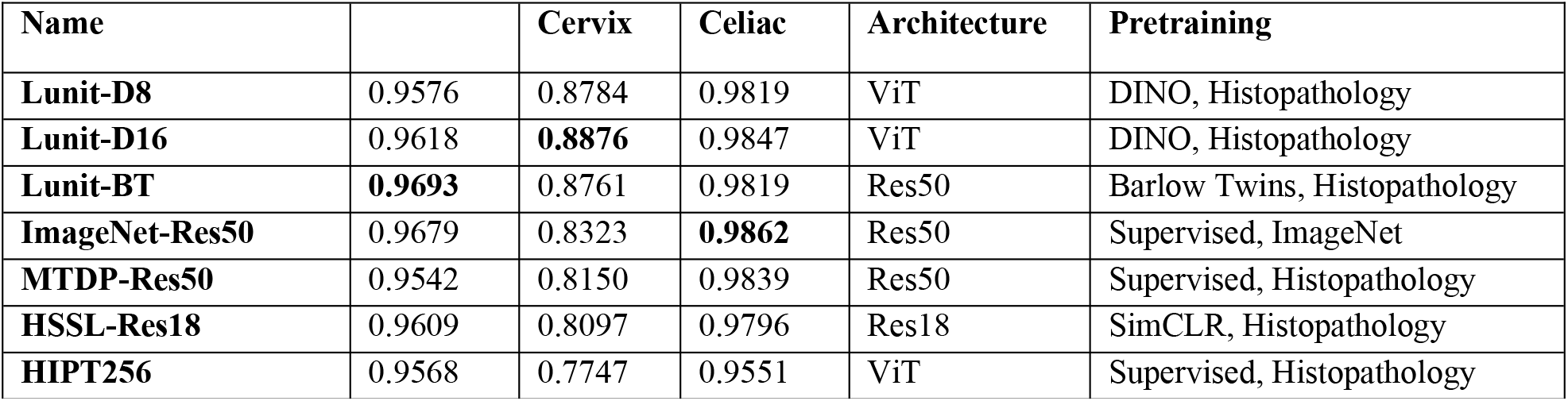
Best test performance (average AUC) of the evaluated encoders; the highest respective score is marked in bold.

| Name |  | Cervix | Celiac | Architecture | Pretraining |
| --- | --- | --- | --- | --- | --- |
| Lunit-D8 | 0.9576 | 0.8784 | 0.9819 | ViT | DINO, Histopathology |
| Lunit-D16 | 0.9618 | **0.8876** | 0.9847 | ViT | DINO, Histopathology |
| Lunit-BT | **0.9693** | 0.8761 | 0.9819 | Res50 | Barlow Twins, Histopathology |
| ImageNet-Res50 | 0.9679 | 0.8323 | **0.9862** | Res50 | Supervised, ImageNet |
| MTDP-Res50 | 0.9542 | 0.8150 | 0.9839 | Res50 | Supervised, Histopathology |
| HSSL-Res18 | 0.9609 | 0.8097 | 0.9796 | Res18 | SimCLR, Histopathology |
| HIPT256 | 0.9568 | 0.7747 | 0.9551 | ViT | Supervised, Histopathology |

### Assessing biopsy diagnoses

A panel of pathologists scored biopsies in two independent sets of cases for colon and two sets for cervix. The same cases were also scored with the proposed NIC-A framework equipped with the best encoder for each application. Figures 2 and 3 depict the results on the evaluation sets of colon and cervix cases, respectively, with data from Radboudumc (Figure 2a, 3a) and ASP (Figure 2c, 3c). The classes benign, LGD, HGD and cancer are depicted in green, yellow, orange and red, respectively, whereas slides that were deemed as non-diagnosable are depicted in gray. Cases are sorted based on the consensus label, considered here as the reference standard. Subfigures (b) and (d) depict the confusion matrices comparing NIC-A performance versus the pathologists’ consensus scores, whereas subfigures (e) and (f) depict quadratic weighted kappa (QWK) scores between the pathologists including the original report, and NIC-A models (excluding the unscored slides). Kappa-scores were weighted non-linearly, with the order benign (1), LGD (2), HGD (3) and cancer (4).

**Figure 3:**
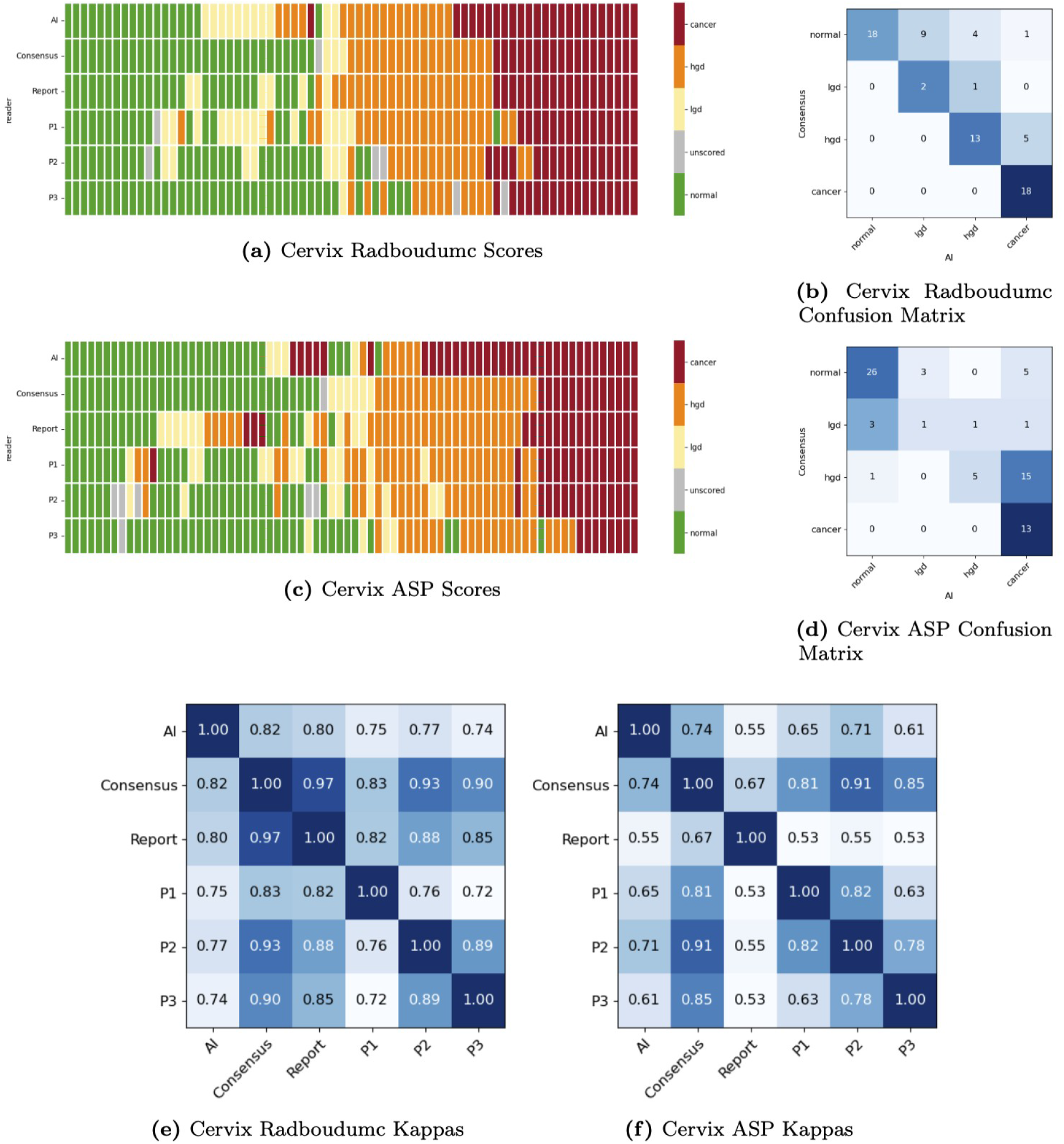
Cervix reader study evaluation results. Slide scores (a,c), confusion matrices when compared to the consensus (b,d) and quadratic kappas for the inter-observer agreement (e,f) for the Radboudumc and ASP cohorts, respectively.

### Colonic specimens evaluation

In the Radboudumc evaluation cohort, NIC-A-colon reaches a substantial agreement with pathologists’ consensus (QWK=0.75) and high agreement with the diagnostic reports (QWK=0.82). The agreement between the pathologists is also very high, with a range between 0.83 and 0.94. On the Catania evaluation cohort, the agreement between the AI and consensus is moderate (QWK= 0.48), while the agreement with the reports is substantial (QWK=0.63). The agreement between the pathologists is in a range of substantial to very high (QKW=0.63-0.91).

NIC-A-colon achieves a high accuracy in differentiating benign from non-benign cases. Compared to the consensus, NIC-A-colon solely downgraded one case in the ASP cohort from LGD to normal, which was reported by one of the pathologists as being a sessile serrated lesion. Two false positive classifications occurred in each cohort: two normal Radboudumc cases were misclassified as LGD (each also diagnosed as LGD by one pathologist) and two normal ASP cases were misclassified as cancer (one submucosal leiomyoma and one case with acute inflammation with ulcus and granulation tissue that was not scored by two pathologists). Overall, the performance of NIC-A-colon was shown to be highly accurate, comparable to that of pathologists. This is also reflected by the performance in terms of ROC curves (see Fig. 3 top supplementary material), reaching 100% sensitivity with less than 30% false-positive rate. Although NIC-A-colon was trained on Radboudumc cases only, we observed similar performance on external evaluation data from Radboudumc and ASP, showing the consistency of the models ability to diagnose unseen cases from both in-domain and out-of-domain data.

### Cervical specimens evaluation

In the Radboudumc evaluation cohort, NIC-A-cervix shows an almost perfect agreement with both the pathologists’ consensus (QWK=0.82) and the diagnostic reports (QWK=0.82). Notably, inter-pathologist agreement on this cohort ranged from 0.72 to 0.89, which is lower than the one obtained for the colon study (0.83-0.94), indicating the higher complexity of the task in cervix tissue grading. As observed in the colon study, agreement on the ASP cohort was again lower, with NIC-A-cervix achieving a QWK=0.74 with the consensus and QWK=0.55 with the reports. Similarly, inter-pathologist agreement for ASP was between 0.63 and 0.82.

NIC-A-cervix shows a similar over-grading trend as its colon counterpart, misclassifying fourteen Radboudumc and seven ASP benign cases as abnormal (thirteen of them were also scored as abnormal (ten LGD and three HGD) by at least one pathologist). While NIC-A-cervix correctly identified all Radboudumc (pre)-malignant cases, four abnormal ASP cases, all of them squamous cell lesions, were missed. In general, as in the colon study, we observed that NIC-A-cervix approached the pathologists performance, particularly on the Radboudumc cohort, where, again, 100% sensitivity with less than 30% false positive rate could be reached (see Fig. 3 bottom supplementary material).

### Duodenal specimen evaluation

Given the binary nature of this task and the difference in diagnosis across countries, such as the use of immunohistochemistry, we did not design a reader study for this application and instead relied on the original report as the reference standard.

On the Radboudumc evaluation cohort, the model reaches an AUC of 0.947 and a kappa score of 0.52 using the labels from the reports as reference standard. The ROC curve and confusion matrix are shown in Fig. 4 on the left and right, respectively. Out of 100 cases, NIC-A-celiac missed three cases with CD and had 21 false positives (normal cases misclassified as CD).

**Figure 4:**
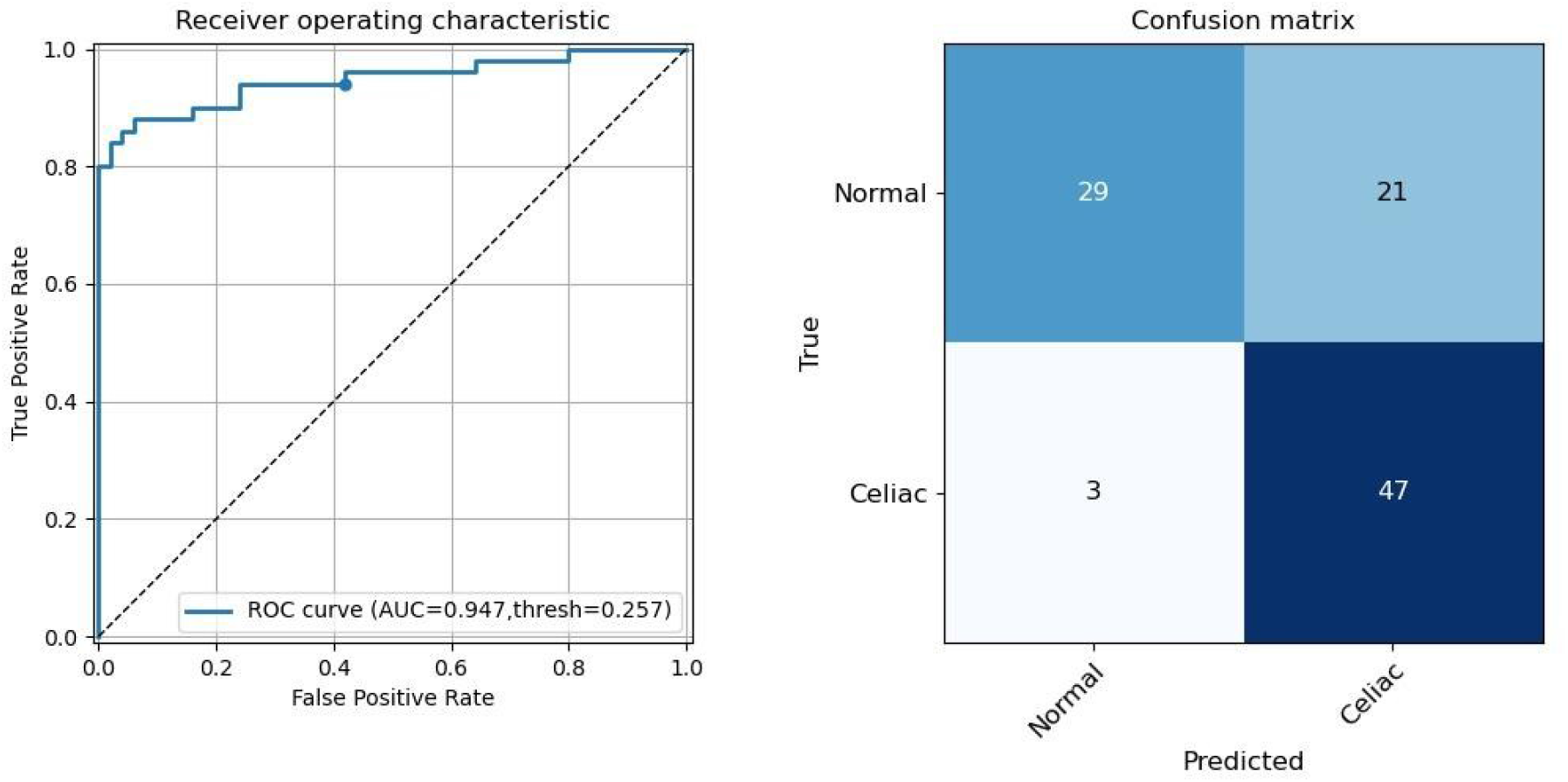
Celiac evaluation results on the Radboudumc cohort: Receiver Operating Characteristic with the cut-off threshold marked with a dot on the left and corresponding confusion matrix on the right.

### Interpretability of the model predictions

The attention-based classification approach allows one to visually inspect the parts of the slide responsible for the model predictions. The attention visualizations for selected slides can be found in Fig. 4-6 in the Supplementary Material. Overall, we observed that the model’s attention focuses on abnormal tissue.

## Discussion

This study investigated the application of a deep learning framework, NIC-A, for pre-screening and subtyping biopsies from colorectal, cervical, and duodenal tissues. We demonstrated the ability of our framework to be trained to address subtyping tasks across different tissue types. Furthermore, for colon and cervix we focused specifically on biopsies derived from population screening programs, reflecting real-world clinical scenarios. The performance of NIC-A was benchmarked against pathologist consensus diagnoses on colorectal and cervical biopsies, providing valuable insights into the model’s agreement with expert opinion.

Our AI models achieved high predictive performances for colonic and cervical specimens subtyping, reaching the diagnostic accuracy of the pathologists. The performance of celiac disease detection was also high. Remarkably, this was achieved by training the model solely using block-level labels without the need of manually annotating slides, also indicating the effectiveness of the proposed packing approach. Furthermore, the performance of our method showed robustness (see Figure 2, 3) to variation in data coming from a different pathology laboratory (ASP) from a different country, scanned with a different scanner from the data source used for training (Radboudumc).

In general, we observed that NIC-A-colon tends to over-grade cases (classifying LGD cases as HGD and some HGD cases as invasive cancer). This tendency towards over-grading could be potentially used to benefit pre-screening colon polyps and biopsies. Due to the demonstrated high accuracy in identifying benign cases, this approach could hold potential value for being used in case triaging in clinical settings, ensuring for example that no abnormal cases are missed during pre-screening.

Similar application scenarios are possible for NIC-A-cervix, which also demonstrated a high sensitivity, particularly on Radboudumc cases. This is a significant strength, as high sensitivity in histology diagnostics minimizes the risk of false negatives (missed abnormal cases), which is especially important for conization specimens, where numerous histology sections require careful examination. A high-sensitivity AI like NIC-A-cervix could be a useful tool for pathologists, allowing them to focus on potentially abnormal sections identified by the model.

Despite the promising results, cervical subtyping presented more challenges to the algorithm than colon subtyping. The reader study for cervix revealed higher inter-observer disagreement among pathologists compared to the colon cohort. This suggests inherent complexities in classifying cervical lesions, even for trained specialists. Additionally, a larger number of cervical specimens were deemed not scorable due to limitations of H&E staining alone. This highlights the potential need for incorporating additional information, like immunohistochemistry (IHC) data, for definitive diagnosis in some cases. Furthermore, the training dataset for cervix subtyping was substantially smaller compared to colon subtyping.

From a technical perspective, the evaluation of the different encoders (pre-training methods) did not yield a single winning method. For colon subtyping, the difference between all models is relatively small, suggesting that the choice of a specific pre-training method is less important if ample training data is available. Conversely, for cervix subtyping with limited data, the three Lunit models have very similar performance while outperforming the other encoders by a considerable margin. This indicates that the amount of the training data and encompassing training procedure are more important than the particular architecture, since the other encoders were trained in similar ways. Interestingly, for celiac detection, the ImageNet-pretrained model had the highest performance, albeit by a small margin. Overall, the choice of the encoder can be considered to be a hyper-parameter setting, to be tuned on the task at hand.

Our deep learning models achieve promising results for tissue subtyping, trained efficiently using labels at the paraffin-block level. One key strength of our approach is its ability to highlight image regions most relevant to the prediction through attention scores. While this could provide valuable cues to pathologists, it doesn’t encompass all potentially relevant morphological features. Future work could explore methods to improve the interpretability of the attention scores, allowing a more in-depth analysis of the model. Furthermore, incorporating additional data including cancer types and conditions such as serrated lesions will create a more comprehensive pre-screening system. Finally, before applying the models in clinical practice, evaluation on larger independent datasets, especially for celiac disease, would be required, as well as the assessment of their potential impact and optimal way to deploy them in clinical practice via Health Technology Assessment analysis, which is part of our future work.

## Conclusion

In this work, we showed that compression-based, attention-driven deep learning for the analysis of WSIs from tissue blocks can achieve pathologist-level performance when classifying histological subtypes of colorectal and cervical biopsies. Classifiers based on self-supervised pre-training yielded the best results achieving high agreement with pathologists with kappa scores up to 0.75 for colon subtyping and 0.82 for cervix subtyping. The models achieved high ROC-AUC scores and could reach 100% sensitivity with a low false positive rate, provided appropriate calibration. The prediction performance for celiac detection was also promising with a kappa score of 0.52. However, the models are currently limited to specific diagnoses and might miss other clinically relevant conditions. Future work will include more detailed subtyping, e.g. including the serrated polyps for colon and differentiating between squamous cell carcinoma and adenocarcinoma in the cervix. These advancements may aid pathologists in achieving less biased and more efficient diagnoses in the future.

## Acknowledgment

This project was funded by the European Union’s Horizon 2020 research and innovation programme under grant agreement No 825292 (ExaMode, http://www.examode.eu/).

## Data availability

Due to data agreement limitations, the patient data used to develop the models cannot be shared. The algorithm source code is available at:

https://github.com/DIAGNijmegen/pathology-whole-slide-packer

https://github.com/DIAGNijmegen/pathology-whole-slide-learning

We made the algorithms for the colorectal and cervical subtyping as well as celiac classification available on the Grand Challenge platform:

https://grand-challenge.org/algorithms/colon-subtyping

https://grand-challenge.org/algorithms/cervix-subtyping

https://grand-challenge.org/algorithms/celiac-classification

## Contributors

FC and WA designed this study. WA performed the experiments, analyzed the results and wrote the manuscript. CvdP, IN, EM, MC, CB, and LA scored the slides in the reader studies. FC and JvdL were involved in supervising the work. All authors reviewed the manuscript and agreed with its contents.

## Ethics Approval and Consent to Participate

The use of the patient data for the study was approved by the Committee on Research Involving Human Subjects of the Radboud University Medical Center (dossier number 2018-4764) and by the Ethical Committee of the Cannizzaro Hospital (approval number 4428, 12/12/2018). During their cancer treatment, patients were informed that left-over tissue could be used for research. Those, who did not authorize the use of their data for scientific research (i.e., decided to opt-out) were not included in the study. This research was performed following the Declaration of Helsinki.

