## Supplementary Material for "Attention-based whole-slide image compression achieves pathologist-level pre-screening of multi-organ routine histopathology biopsies"

### 1. Reader Studies

Suppl. Fig. 1 shows an example of the graphical interface for the reader studies.

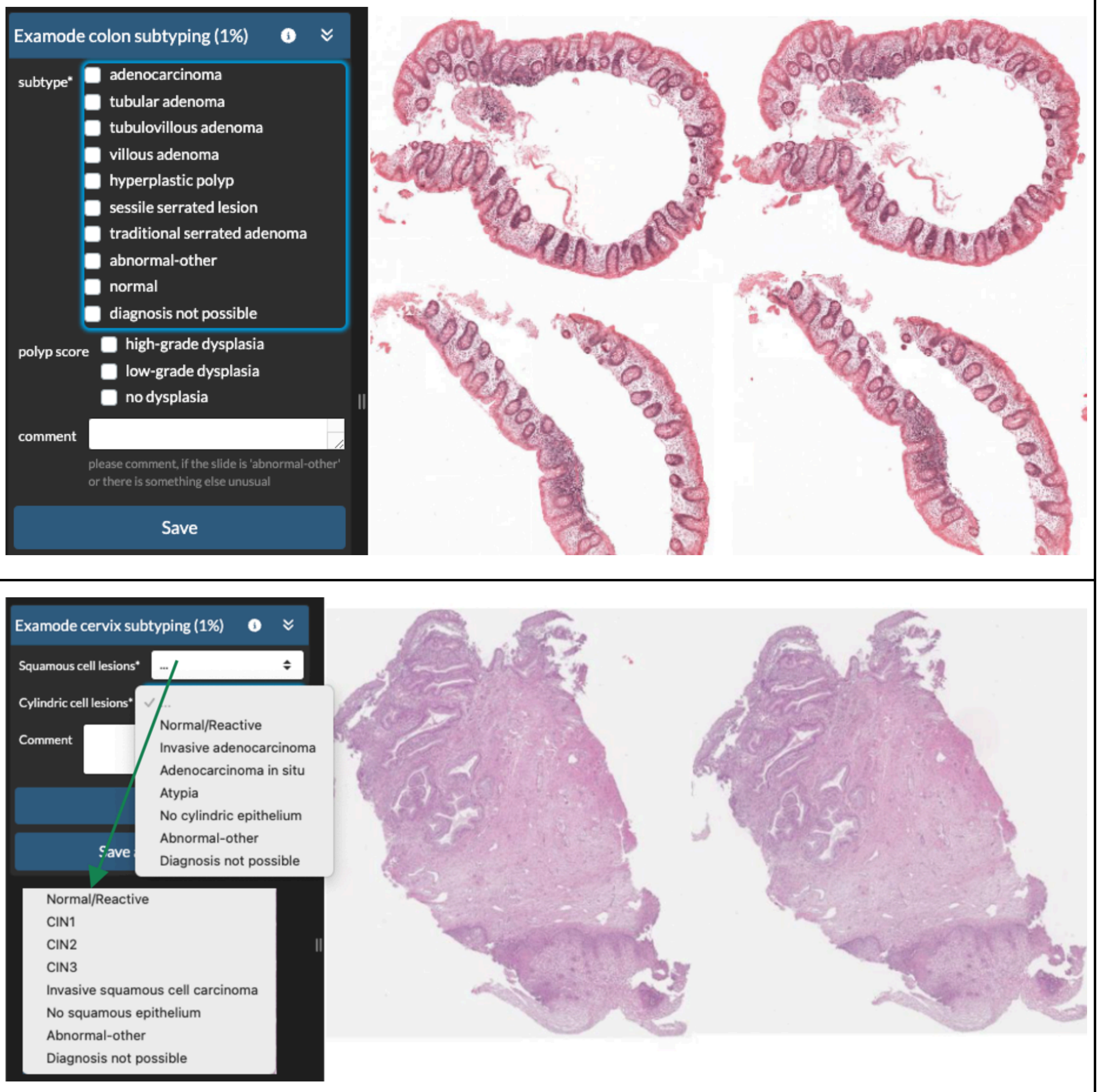


Suppl. Fig. 1: Colon and Cervix reader study examples on the top and bottom, respectively (screenshots taken from https://grand-challenge.org)

### 2. Attention Based Neural Image Compression Classifier Architecture

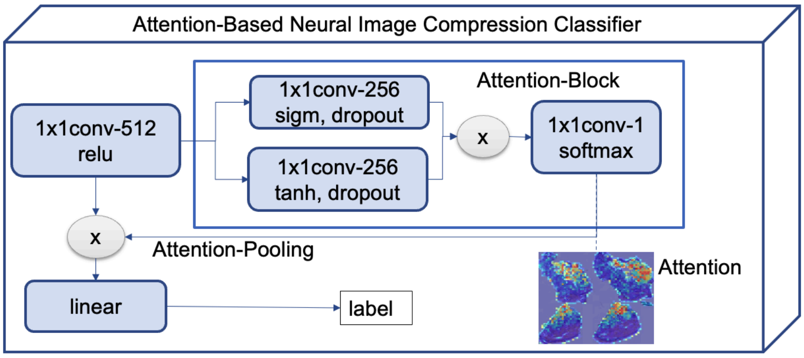


Suppl. Fig. 2: Architecture of the Attention Based Neural Image Compression Classifier. The first layer reduces the number of channels of the encoded slide to 512. Then, the attention scores are computed: features from two layers with 256 neurons are combined via dot-product, followed by a layer creating a single score for each compressed patch. The softmax ensures that the scores are in range [0,1]. The scores are then multiplied with the vectors from the first layer and the results are added via a matrix multiplication ('Attention-Pooling'). This results in a single vector of size 512 which is then converted to a single prediction by the final linear layer.

### Multi-label Classification/Per-Subtype Analysis

Our AI models were trained for tissue subtyping of colon and cervix as a multi-label classification problem. The main analysis focuses on the worst-case scenario, where only the most hazardous subtype is considered allowing a multi-class analysis (i.e. if cancer is present, presence of high grade dysplasia is disregarded). However, the analysis can be further extended to evaluate the model's performance for each individual subtype using a one-vs-all approach. In this approach, the model is evaluated for its ability to classify each subtype as present or absent, considering all other subtypes as negative. Suppl. Fig. 3 depicts the receiver operating characteristic (ROC) curves for each tissue subtype on the colon and cervical datasets, along with the placement of the pathologist diagnoses for comparison.

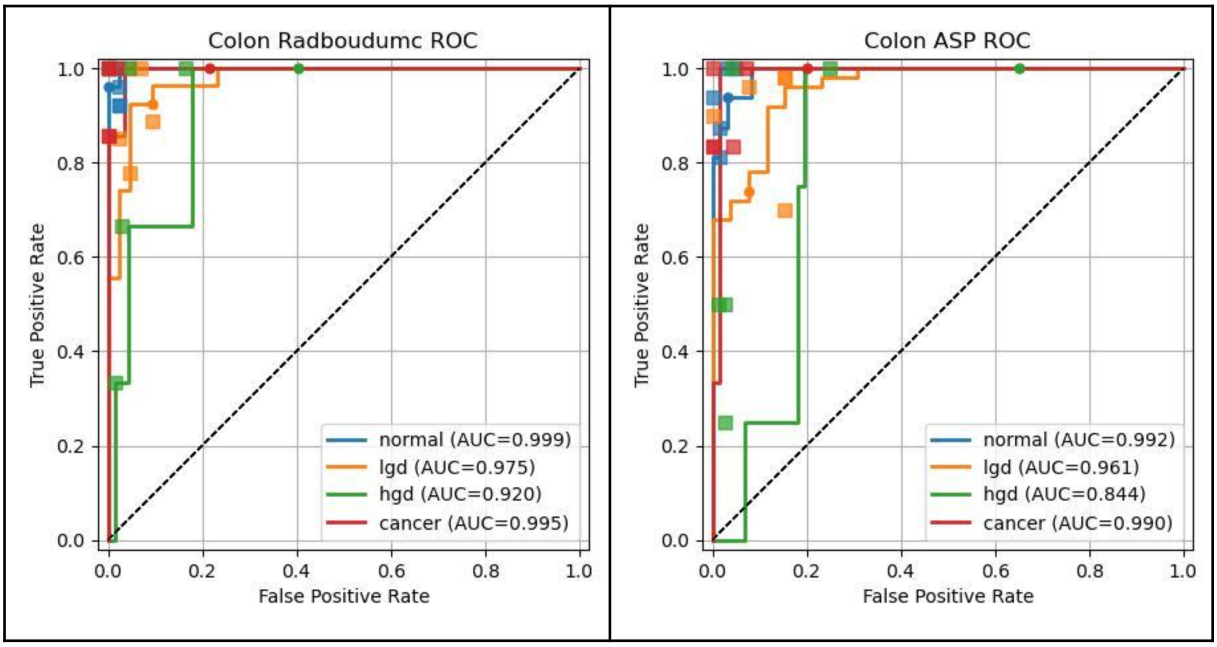

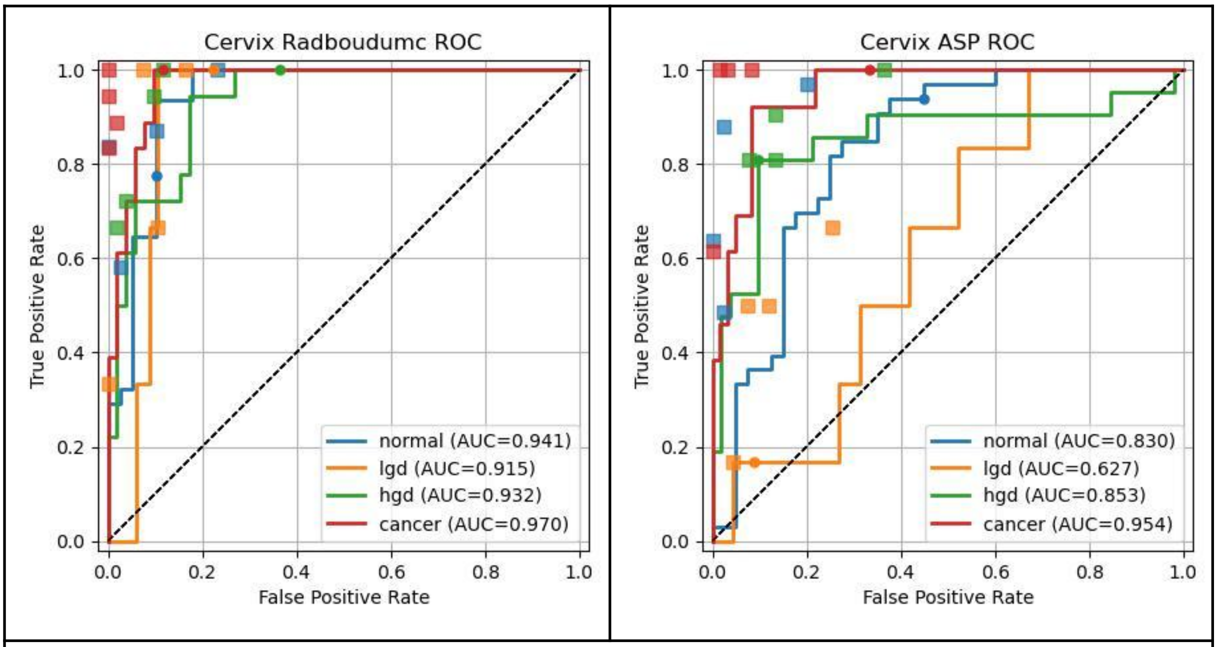


Suppl. Fig. 3: ROC curves of the AI models on each tissue subtype. The AI-cutoff thresholds are marked with dots. The pathologist results are depicted with squares in the color of the corresponding tissue type.

### 3. Attention Maps

An important part of the NIC-A architecture are the attention scores, which are used to weight the patch-encodings before classification. In the following figures we visualize the attention scores for several whole slide images in the evaluation sets via heatmaps (green: low attention, yellow/orange: moderate attention, red: high attention). The attention visualizations highlight diagnostically relevant areas, suggesting that with better calibration the sensitivity of the model could be improved.

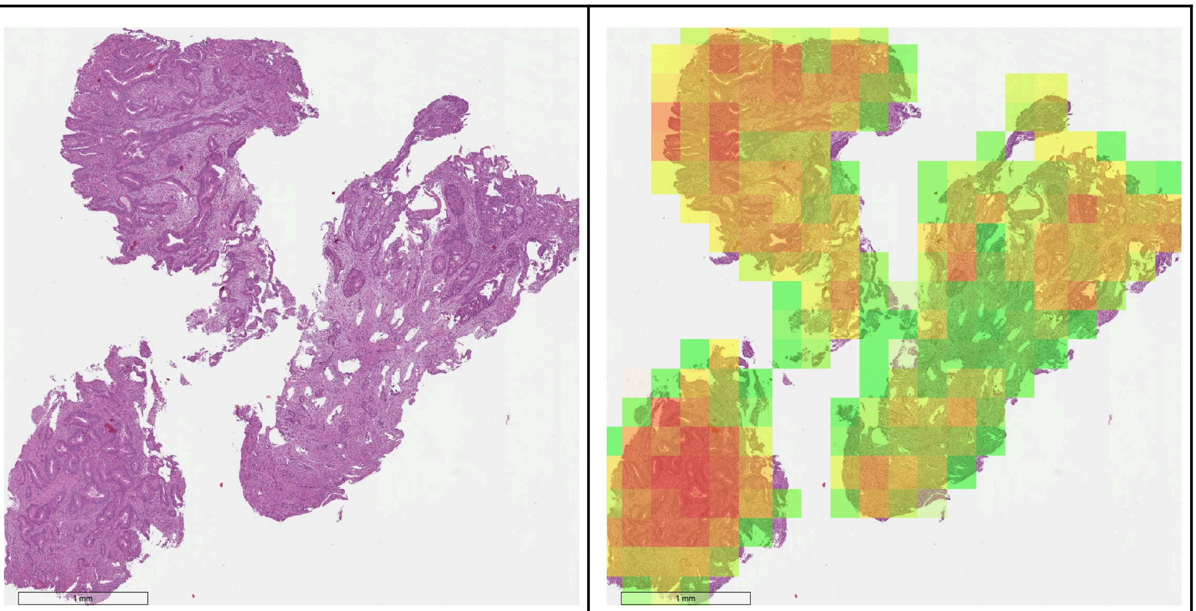

Suppl. Fig 4A. Correctly classified colon cancer example from the ASP cohort.

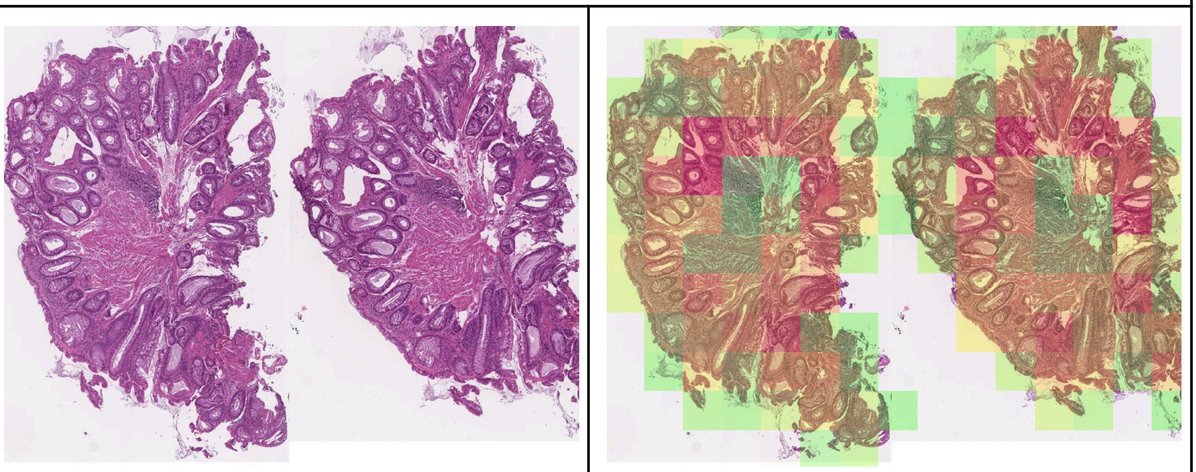

Suppl. Fig. 4b: Incorrectly classified colon lgd example from ASP. However, the model correctly identified dysplastic areas.

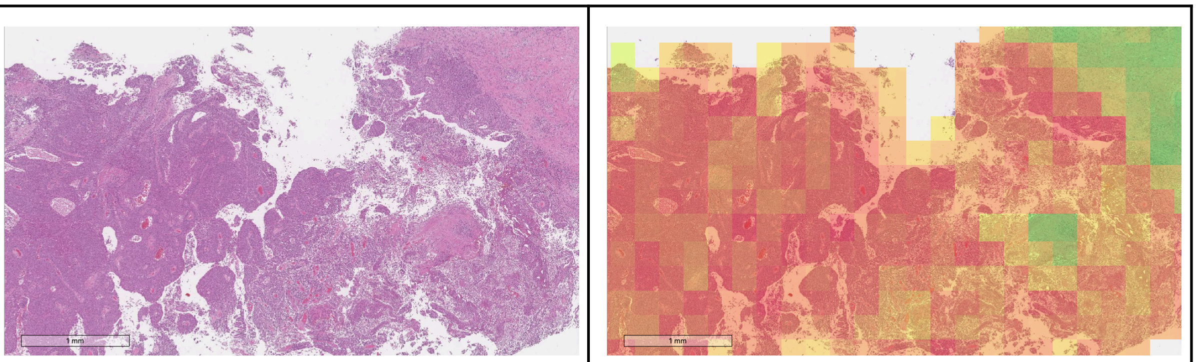

Suppl. Fig. 5a: Correctly classified invasive adenocarcinoma (cylindrical cell lesion) from ASP.

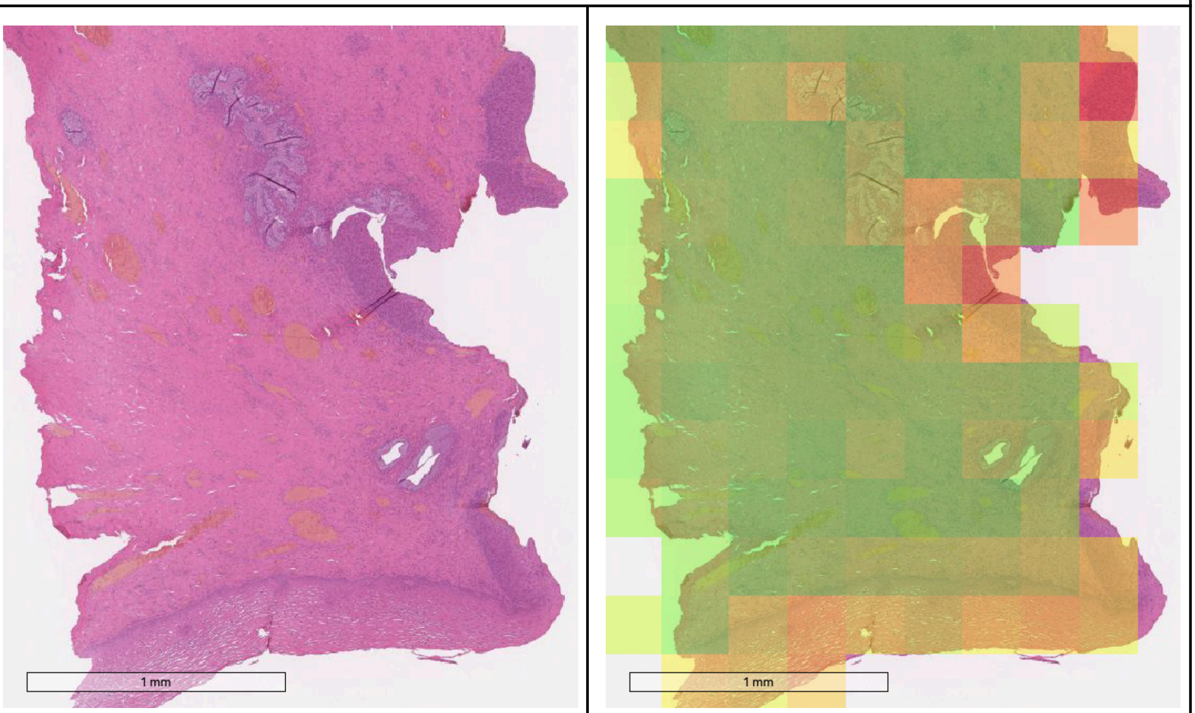


Suppl. Fig. 5b: Incorrectly classified cervical high grade squamous cell lesion from ASP. However, the model correctly identified the lesion area.

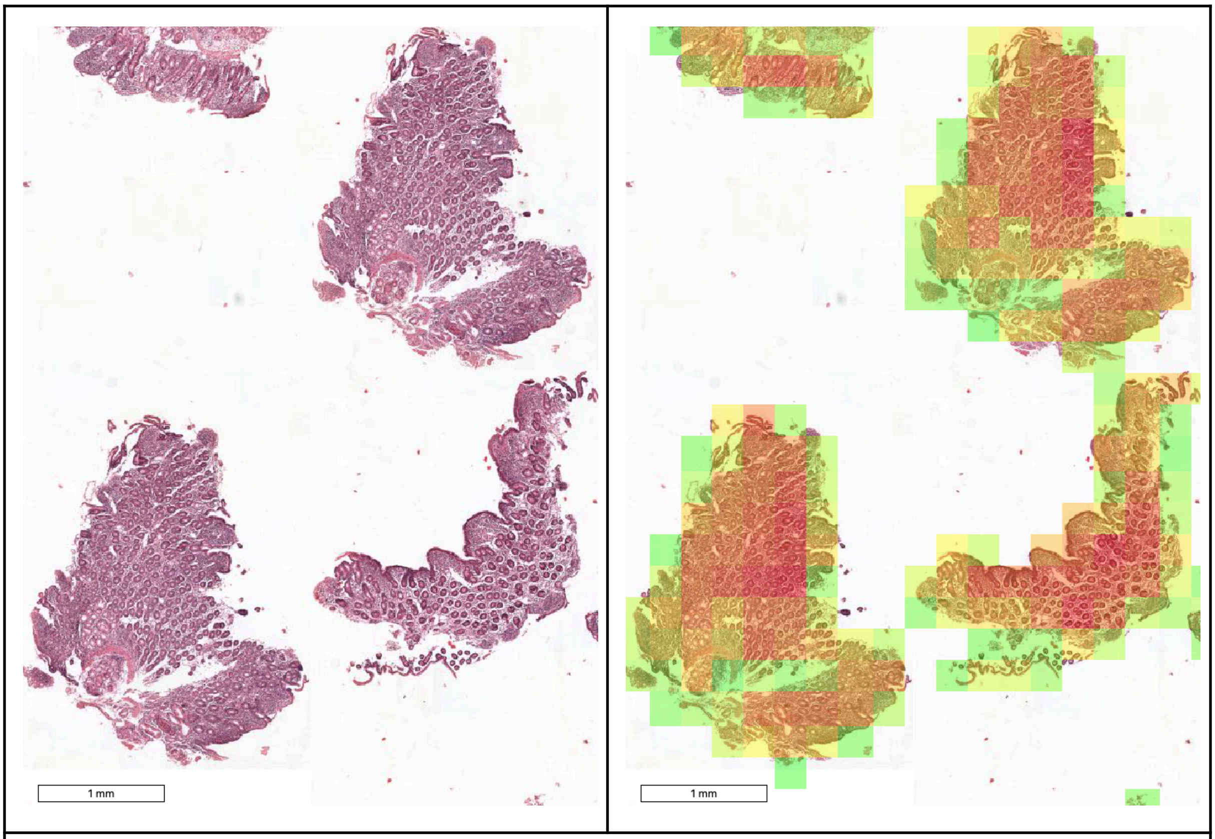


Suppl. Fig. 6A: Correctly classified celiac disease sample from the Radboudumc cohort.

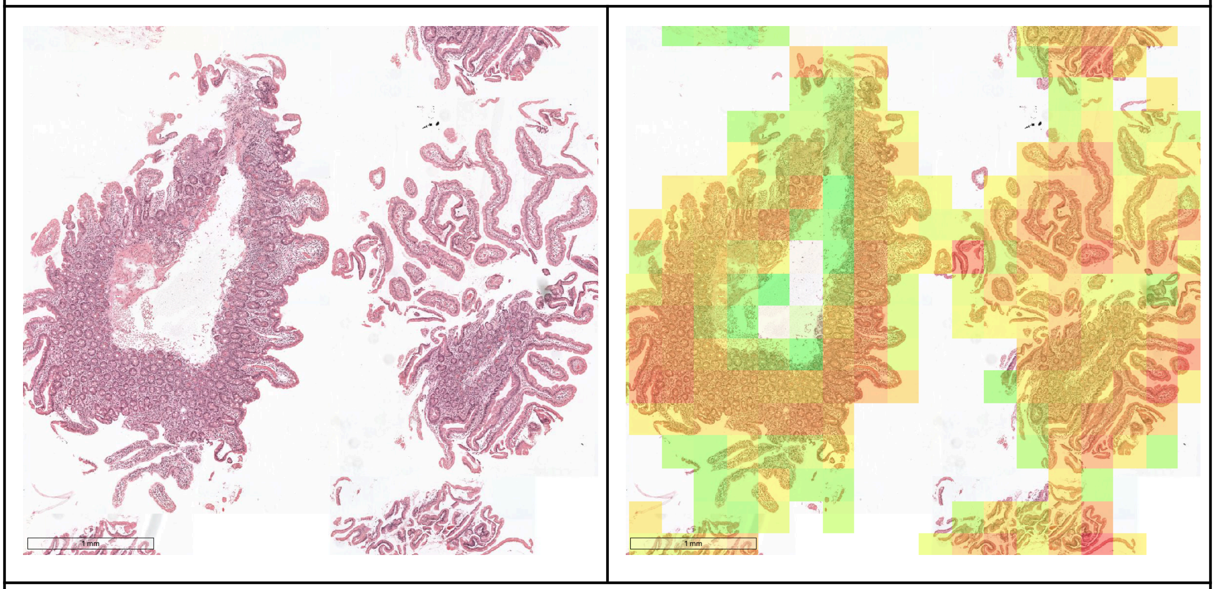

Suppl. Fig. 6B: Incorrectly classified celiac disease sample from the Radboudumc cohort.
